# Diagnosis of spine pseudoarthrosis based on the biomechanical properties of bone

**DOI:** 10.1101/2024.01.06.23300551

**Authors:** John A Hipp, Mark M. Mikhael, Charles A Reitman, Zorica Buser, Vikas V. Patel, Christopher D. Chaput, Gary Ghiselli, John DeVine, Sigurd Berven, Pierce Nunley, Trevor F Grieco

## Abstract

**Background:** Cervical spine fusion, commonly performed with generally favorable outcomes, may result in postsurgical symptoms requiring further investigation and treatment. Anterior cervical discectomy and fusion (ACDF) aims to decompress neural structures, stabilize motion segments, eliminate intervertebral motion, and promote bridging bone formation. Failure to form bridging bone may result in persistent symptoms or symptomatic pseudoarthrosis. Traditional diagnosis involves computerized tomography to detect bridging bone and/or flexion-extension radiographs to assess whether segmental motion is above specific thresholds. This paper proposes a new biomechanically based diagnostic approach to address limitations in traditional diagnostic methods. The scientific basis of this approach is that bridging bone cannot occur if the strain is greater than the failure strain of the bone.

**Methods:** Fully automated methods were used to measure disc space strains. Errors in strain measurements were assessed from simulated radiographs. Measurement error combined with the reported failure strain of trabecular bone led to a proposed strain threshold for pseudoarthrosis diagnosis post-ACDF surgery. A reanalysis of previously reported flexion-extension radiographs for asymptomatic volunteers was used to assess whether flexion-extension radiographs, in the absence of fusion surgery, can be expected to provide sufficient stress on motion segments to allow for reliable strain-based fusion assessment. The sensitivity and specificity of strain- and rotation-based pseudoarthrosis diagnosis were assessed by reanalysis of previously reported post-ACDF flexion-extension radiographs, where intraoperative fusion assessments were also available. Finally, changes in strain over time were explored through the use of 9,869 flexion-extension radiographs obtained 6 weeks to 84 months post-ACDF surgery from 1,369 patients.

**Results:** The estimated error in measuring disc space strain from radiographs was approximately 3%, and the reported failure strain of bridging bone was less than 2.5%. On that basis, a 5% strain threshold is proposed for pseudoarthrosis diagnosis. Good-quality flexion-extension radiographs can be expected to stress the spine sufficiently to facilitate strain-based diagnosis of pseudoarthrosis. Reanalysis of a study in which intraoperative fusion assessments were available revealed 67% sensitivity and 82% specificity for strain-based diagnosis of pseudoarthrosis, which is comparable to rotation-based diagnosis. Analysis of post-ACDF flexion-extension radiographs revealed rapid strain reduction for up to 24 months, followed by a slower decrease for up to 84 months. When rotation is less than 2 degrees, the strain-based diagnosis differs from the rotation-based diagnosis in approximately 14% of the cases.

**Discussion:** Steps for standardizing strain-based diagnosis of pseudoarthrosis are proposed based on the failure strain of bone, measurement error, and retrospective data. These steps include obtaining high-quality flexion-extension studies, the application of proposed diagnostic thresholds, and the use of image stabilization for conclusive diagnosis, especially when motion is near thresholds. The necessity for an accurate diagnosis with minimal radiation exposure underscores the need for further optimization and standardization in diagnosing pseudoarthrosis following ACDF surgery.

## Introduction

### General background

Pseudoarthrosis is an important cause of failed cervical spine surgery and an important reason for reoperation.(1–5) Diagnosis of the specific cause(s) for symptoms post-fusion surgery is challenging and complicated in part by the observation that pseudoarthrosis in the cervical spine can be asymptomatic and that not all patients with pseudoarthrosis need to be revised.(5–8) Nevertheless, one possible reason for post-fusion symptoms is failure to achieve the technical goal of fusion, which is elimination of intervertebral motion through the formation of bone bridging between vertebrae. Accurate measurement and interpretation of intervertebral motion are essential for accurate differential diagnosis of patients with pain after cervical spine fusion.

The following discussion focuses on surgery intended to achieve fusion across the disc space between vertebral bodies in the cervical spine. Assessing posterior fusions intended to achieve fusion only across the posterior elements of the spine is a different challenge. In the following analysis, an interbody bridging bone was assumed to be required for technically successful anterior fusion. Although the exact tissue composition that occurs with asymptomatic pseudoarthrosis is poorly documented, it may be possible that bridging fibrocartilaginous tissue in the disc space(9) provides sufficient stability to relieve symptoms. Nevertheless, the following paradigm for the diagnosis of pseudoarthrosis assumes that the technical goal is bridging bone and not sufficiently stiff nonmineralized tissues. The focus will be on anterior cervical spine fusion, although the basic concepts are directly applicable to lumbar interbody spine fusion.

No clinically practical fusion assessment protocol based on radiographs or CT exams has ever been proven to be highly reliable, as summarized in multiple review papers (10–22). Reviews typically conclude that the best currently available fusion assessment option is subjective assessment of bridging bone and the lack of a radiolucent line from thin-slice CT exams combined with intervertebral motion measurements, although the reviews acknowledge certain limitations.

The proliferation of numerous review papers underscores the broad interest in addressing this issue. However, the lack of standardized tests for assessing spine fusion success in everyday clinical practice indicates that a definitive solution to this problem has yet to be identified. Enhanced communication among physicians, research into optimal treatment strategies and technologies, and improved patient outcomes could all benefit from the implementation of validated and standardized diagnostic tests.

As a supplement to existing reviews(10–22), appendix 1 provides a critique of previously described and applied approaches to using fusion assessment via intervertebral motion measured from flexion–extension radiographs or bridging bone assessment via CT exams.

The essential goal of fusion is to eliminate movement between adjacent vertebrae. While many studies use intraoperative evaluation of intervertebral movement as the reference standard for fusion assessment, this method lacks adequate validation. (12, 23–31) Relying on subjective, invasive intraoperative evaluations in routine clinical practice is not ideal because the applied force and the subjective assessment of motion are difficult to standardize among surgeons. The force applied by a surgeon is likely minor compared to the force exerted by patients when trying to flex and extend their spine to the maximum. Therefore, an innovative approach for assessing spine fusion may be essential for achieving a universally accepted standard diagnostic test for pseudoarthrosis in both research and clinical settings.

It is reasonable to hypothesize that noninvasive intervertebral motion measurements are reliable for assessing technical fusion success if two conditions are satisfied:

1. Intervertebral motion (IVM) measurements are accurately derived from two spinal images taken under significantly different loading conditions to ensure detectable IVM if the level is not fused.
2. A biomechanically rational approach was used to interpret the measurements.

### A novel approach to the diagnosis of pseudoarthrosis

Technically successful spine fusion requires bone bridging between adjacent vertebrae. Bone will only tolerate a small amount of strain before it fractures (32). Strain is the change in length caused by a change in loading divided by the initial length. Bridging bone **CAN NOT** occur between endplates if the strain exceeds the failure strain of the bone. This biomechanical fact allows for a definitive fusion assessment paradigm. Strain has previously been studied and used for spine fusion assessment in research studies. (54, 55) Strain-based diagnosis of pseudoarthrosis should optimally be based on an understanding of the type of bone that connects vertebrae.

Unfortunately, the mechanical properties of the tissues that exist between vertebral endplates from the time of surgery through definitive solid bridging have been poorly documented. Bone healing occurs in well-defined stages, including inflammatory, regenerative and remodeling stages. The remodeling phase of bone healing leads to trabecular bone within the fusion mass.(31, 33–36) It is reasonable to assume that the bone that bridges between endplates has failure properties similar to those of trabecular bone from vertebral bodies. The yield strains of trabecular bone from different anatomic sites are very similar(37), which provides confidence in the application of existing vertebral trabecular bone failure data to diagnostic tests for spine fusion. In addition, the wealth of knowledge about bone fracture healing(38, 39) has relevance to the challenge of understanding the bone that may bridge vertebrae. Early-stage bone healing may tolerate somewhat greater amounts of strain than true bridging bone(33, 40). Therefore, the strain thresholds used in a diagnostic test ideally depend on the relevant stage of bone healing.

As tested in a laboratory, a bone specimen initially behaves elastically as the applied load increases, then exhibits nonlinear behavior and finally fractures.(41) This behavior is well documented and repeatable.(42) The failure strain of trabecular bone samples (human and animal) in laboratory studies is < 2.5%.(32, 43–47) Note that some laboratory studies report a higher failure strain, but this difference can be explained by end artefacts during testing.(43) The ultimate strain (strain at maximum deformation right before failure) of trabecular bone from vertebral bodies has been documented to be 1.59±0.33.(43) Assuming a normal distribution, the upper limit of the 95% confidence interval would be 2.24%. Accounting for errors in the strain measurements of approximately 3% (as discussed in Appendix 2), a threshold level of >5% strain is a reasonable threshold for a diagnostic test for pseudarthroses.

With accurate measurements of disc space widening, apparent strain can be measured in clinical practice from two images of the spine subjected to two different loading conditions (e.g., flexion versus extension). Apparent tensile strain in the disc space between vertebrae can be measured as the change in disc space width divided by the smallest disc height found in the two images. The word “apparent” is used since strain in individual bone trabeculae is not being measured, and trabeculae comprising bridging bone do not always form in a straight line from the endplate to the endplate. (31, 34–36) The “apparent” strain measured between a point on the inferior endplate and a point on the superior endplate may not be equal to the tissue strain occurring in individual spicules of bridging bone.(48) Nevertheless, apparent strain may prove to be an effective metric. Tensile strain is emphasized over compressive strain, assuming that tensile strain is more detrimental to the formation of bridging bone between vertebral endplates.

When applied to spine fusion assessment, apparent strain can be measured between two opposing points on the endplates on either side of the disc space. Strain measurements can be made over a wide range of disc heights. Since tensile strain is calculated by dividing the change in disc space height divided by the smallest initial height, bone bridging across a thin disc space will tolerate less displacement between endplates than will a thick disc space(39), which is what is accounted for in strain-based analysis. For example, the maximum strain that occurs between endplates at the anterior most aspect of a disc space for a specific amount of rotation (e.g., 5 degrees) will be much greater when the disc space is 1 mm thick versus 10 mm thick. This is a biomechanical flaw in using rotation as the metric to assess fusions that are overcome by a strain-based fusion assessment.

Strain-based fusion assessment requires accurate measurements of the change in disc space height after fusion surgery. Accurate measurements of IVM motion are currently possible with computer-assisted methods (49–52). An additional advantage of a strain-based approach to fusion assessment is that variability in image magnification will not compromise assessment accuracy since strain is dimensionless. It would **not** be necessary to place a calibration marker in the radiographic field to accurately determine image scaling. Nevertheless, it would be necessary to assure that the magnification is the same for both flexion and extension images or that the method for intervertebral motion measurement compensates for any differences in magnification.

Bono et al.(53, 54) investigated the phenomenon of partial bridging (e.g., anterior) that can allow motion elsewhere (e.g., posterior) by elastic bending of the partial bridging. To account for that possibility, apparent tensile strain could be calculated at multiple points across the disc space (Figure 1). At each location, the shortest distance between two images serves as the denominator, while the distance change is the numerator. This approach can be easily automated with computer-aided technology. If the level is loaded to produce more than 5% strain across the entire disc space (assuming that it is not fused) and if the strain exceeds 5% at every point, then the level is conclusively not fused. If the strain at only some points is < 5%, then the level may be partially fused, and a strain > 5% at other locations is allowed by elastic bending of the bone in the portion of the disc with bridging bone (53, 54). Bono et al recognized the need to use a fusion assessment methodology that accounts for the location of bridging bone (54). Measuring strain at multiple locations across the disc space provides a methodology for measuring partial fusion.

**Figure 1:**
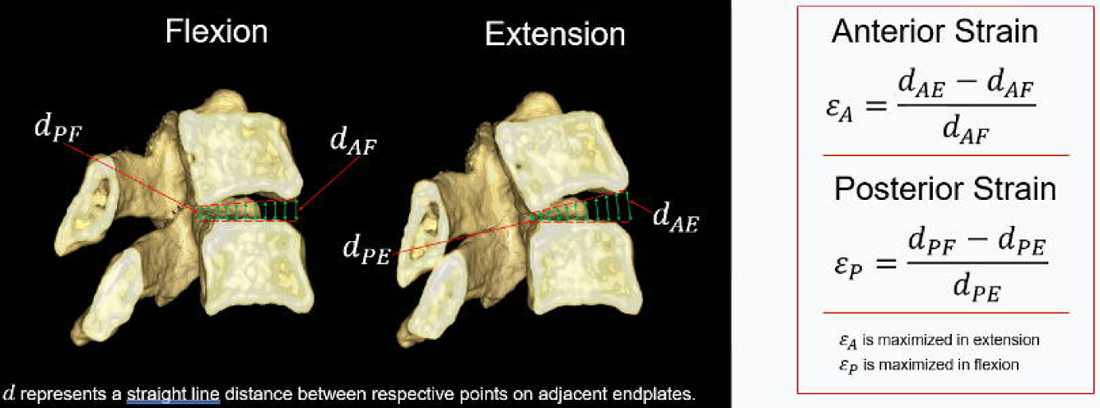
Illustration of how the lines connecting points on the endplates can be used to calculate strain at multiple points across a disc space. This can be done both at the level where fusion surgery had been performed but also at adjacent levels so that the adjacent levels could be used to assess whether the fusion surgery level had been adequately loaded.

## Methods

Several experiments were completed to help better assess the potential of strain-based diagnosis of pseudoarthrosis. In all the described experiments, vertebral landmarks were automatically placed on the four corners of each vertebral body in paired flexion and extension radiographs and subsequently used to calculate strain (Figure 1). The landmarks were obtained using fully automated and FDA-approved intervertebral motion measurement technology (SpineCAMP^TM^, Medical Metrics, Inc., Houston, TX). However, this technology was not the focus of the study, as artificial intelligence and machine learning methods have been developed for use in identifying vertebral landmarks by many researchers. (60–64) The details of landmark placement for this study have been documented previously(65) and are intended to be comparable to those in prior studies. It is anticipated that the landmark placement used for strain measurements in this study can be replicated by alternative methods, making the data valuable for other investigators utilizing alternative methods for obtaining landmarks. All the data analyses were completed using Microsoft Excel and Stata (StataCorp LLC, College Station, TX).

### Errors in measuring strain from flexion-extension radiographs

As with all measurements used for clinical diagnostic tests, there will be errors in measuring strain from flexion–extension radiographs. One source of error is the radiographic distortion caused by radiographs originating from a point source and passing at various angles through the spine to the image plane. In addition, variability between images in the orientation of the radiograph beam could cause error in disc strain measurements. Quantifying the error requires a reference standard known to be highly accurate. An ex vivo laboratory study is one option, but it is difficult to accurately measure strain within a disc space in the laboratory. A second option is to use images representing flexion-extension radiographs where there is certainly no actual motion between vertebrae. In that experiment, any measured motion was a measurement error. This type of experiment was completed and is described in Appendix 2.

### Disc strains in the absence of fusion

Flexion-extension radiographs should only be used if there is confidence that the spine will be adequately loaded to reliably apply strain-based diagnosis of pseudoarthrosis. To understand the strains across the disc space that occur with upright flexion-extension radiographs, in the absence of fusion, strain was calculated at 11 equally spaced points (Figure 1) along the disc spaces of C2-C3 to C6-C7 from previously collected flexion-extension radiographs of asymptomatic volunteers. The inclusion/exclusion criteria and IRB approval have been previously described.(66) All the volunteers were asked how to maximally flex and extend the device. The distance between each point along the superior endplate of the inferior vertebra and the corresponding point along the inferior endplate of the superior vertebra (Figure 1) was measured during flexion and extension by interpolating between the anterior- and posterior-most landmarks of the endplates. The smallest of these distances was used as the denominator for calculating the percent tensile strain (change in distance divided by the smallest distance * 100).

### Reanalysis of imaging and data from Ghiselli et al.(52)

With the goal of documenting the threshold of strain across a fusion site that might be most sensitive and specific for the diagnosis of pseudoarthrosis, images and data from a previously published study were reanalyzed.(52) In that study, intra-operative assessments of ACDF patients were available, along with flexion-extension radiographs. Apparent strain was measured for 20 levels in 10 patients who were at least 1 year postop. The sensitivity and specificity of the strain measurements for predicting the intraoperative assessment of spine fusion were determined. Motion indicating true nonunion was found at 9/20 of the patients intraoperatively.

### Reduction in the maximum strain over time

Strain across the disc space should theoretically decrease over time after ACDF with remodeling of the bridging bone. An anterior plate is used to provide the initial stability required to promote the formation of bridging bone. It has previously been reported that fusion occurs when the strain across the fusion site is low enough to be compatible with the expected strain of the bridging bone (e.g., < 5%).(67) To determine whether the strain measurements demonstrated a reduction over time to a level consistent with that of mature bridging bone, a pooled analysis of previously collected cervical spine flexion-extension studies was performed. In its role as an imaging core laboratory, Medical Metrics, Inc., has collected and analyzed thousands of cervical spine flexion-extension radiographs from many clinical research studies. Pooled analysis of these images was used to establish internal quality control data and methods. An analysis of disc strain measurements over time was completed with the goal of improving quality control assessments.

Flexion-extension radiographs were obtained at 6 weeks to 84 months postop. There were 9869 flexion-extension radiographs analyzed for 1369 patients. Ninety-three of the patients underwent two-level ACDF, while the remaining patients underwent single-level ACDF. The anterior- and posterior-most strains at the superior and inferior adjacent level disc spaces were also analyzed for reference. The analysis was limited to flexion-extension radiographs where the maximum strain at an adjacent level was > 10%. This was done as a step toward ensuring that the spine was adequately stressed to allow for a reliable strain-based fusion assessment. The analysis focused on the disc space strains at the anterior and posterior most aspects of the disc spaces and not on the possibility of partial bridging since there was no method to verify the existence of partial bridging.

## Results

### Errors in measuring strain from flexion-extension radiographs

Assessment of the errors that can occur in strain measurements made from flexion-extension radiographs (Appendix 2) suggested that when the radiographic projection is similar between images, the amount of error in disc strain measurements is small (< 3% on average). The error is greater when the disc space is narrow since strain is calculated as the change in disc space height at a point divided by the smallest height. However, when the radiographic projection is quite different between images, the error in disc strain measurements can reach 10%, and rotation and translation measurements can have errors as high as 1.8 deg and 4.2%, respectively, in terms of endplate width. Strain- or rotation-based pseudoarthrosis diagnosis needs to be applied with caution when poor-quality radiographs or flexion and extension images are not well matched with respect to radiographic projection. Methods for automatically assessing image quality and rejecting inadequate images have yet to be fully described and validated.

### Disc strains in the absence of fusion

Analysis of disc strains in radiographically normal discs from asymptomatic volunteers revealed large strains in the anterior and posterior aspects of the disc space. Using the logic that this represents the strain across the disc space that would occur in the absence of fusion surgery and that this is the strain that fusion surgery needs to stop, the data support that anterior and posterior strains calculated from flexion–extension radiographs should sufficiently stress the level of fusion surgery (Figure 2). However, strains near the middle of the disc may be too small to allow for the use of strains at adjacent levels to assure that the level of prior fusion surgery was sufficient at those locations (Figure 2). Note that the strains at the anterior and posterior most aspects of the disc space were in accordance with previously reported disc space strains.(68–70)

**Figure 2:**
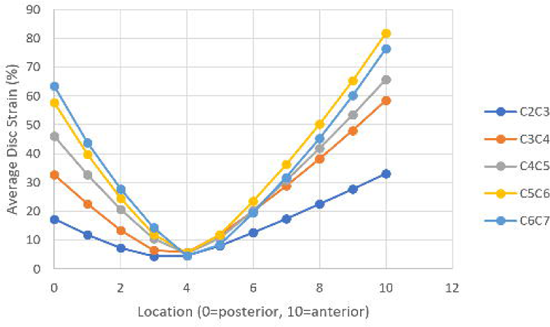
Apparent strain in cervical discs of asymptomatic volunteers between flexion and extension. These data support that in the absence of fusion, with the exception of the central region of the disc, strains would exceed those compatible with bridging bone. This supports that cervical spine flexion-extension studies may sufficiently stress the spine to allow for fusion assessment.

### Reanalysis of imaging and data from Ghiselli et al.(23)

Although the sample size was small (10 patients), reanalysis of flexion-extension studies in which intraoperative assessments of fusion status were also available provided insights into the diagnostic performance of strain- and rotation-based diagnosis of pseudoarthrosis. At all 20 levels that were analyzed, there was at least 10% strain at one of the adjacent levels, providing some assurance that the spines were adequately stressed. The strain threshold that optimized the sensitivity and specificity was 4%. The rotation threshold that optimized the sensitivity and specificity was 1 deg (the same as previously reported(52)). With a 4% threshold, the strain was 67% sensitive and 82% specific for the diagnosis of pseudoarthrosis. Using a one-deg threshold, rotation was 67% sensitive and 90% specific. Based on a comparison of the areas under the ROC curves, the sensitivity and specificity of strain and rotation did not significantly differ (p=0.53). There is currently limited evidence to support prioritizing sensitivity over specificity or vice versa, as there are distinct disadvantages to both under- and over-diagnosing a condition. However, in patients with a high clinical probability, sensitivity may be more useful. Conversely, if the clinical probability is low, then specificity may be more useful. Since the sample in the Ghiselli et al. reanalysis was small, a 5% strain threshold may be a better choice until additional evidence becomes available.

### Reduction in the maximum strain over time

Analysis of 9869 flexion-extension radiographs obtained from 1369 patients at 6 to 84 months postop revealed a more rapid reduction in strain both anteriorly and posteriorly during the first 24 months than after 24 months (Figure 3). The strain was initially greater posteriorly than anteriorly, presumably because the stabilizing effect of the anterior plates was more pronounced anteriorly. By 12 months, the mean anterior strain was nearly 4%, whereas the mean posterior strain did not approach 4% until 24 months. After 24 months, there was a gradual reduction in anterior and posterior strains over time, consistent with the maturation of trabecular bone. There was variability in the rates of fusion between segments, in agreement with prior studies.(71) Note that at 84 months, the average strains are very close to the 2.5% strain that has been reported as the upper limit of the ultimate strain of trabecular bone.(43) Note also that the standard error bars are wider for posterior strains than for anterior strains, supporting greater variability in posterior strains. This is presumably because the anterior plates are generally effective at providing initial stability anteriorly (where the plate is attached). It is reasonable to hypothesize that bone bridging starts anteriorly and progresses posteriorly (with ACDF) as the anterior bridging bone builds on the initial stability provided by the plate.

**Figure 3:**
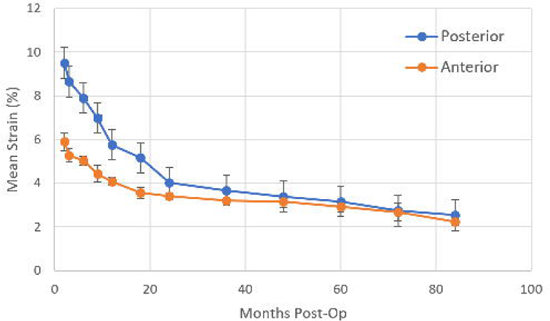
On average, anterior and posterior strains decreased rapidly in the first 1-2 years and then gradually reduced over time. Error bars show the standard error. If strain > 5% any place along the disc is used to classify a level as a pseudoarthrosis, then these data document a 57% fusion rate at 12 months and a 67% fusion rate at 24 months.

Prior to definitive fusion, there is the possibility of stable pseudoarthrosis that is clinically tolerable, as evidenced by the fact that there are cases with strain > 5% out to several years postop. These data also support the observation that some pseudoarthrosis detected one year after ACDF will eventually heal.(72, 73) In addition, these data support the potential for delaying decisions regarding pseudoarthrosis treatment for at least 2 years, as the fusion process may continue to evolve for at least that long in some patients.

The maximum strain (anywhere along the disc but almost always posteriorly after ACDF) and intervertebral rotation were correlated (Figure 4, R^2^ = 0.69, P<0.0001). The levels where the strains were much greater than the rotation were almost always the cases where the disc space was very narrow. This is predictable since strain was calculated as the change in disc space height over the smallest disc height. When the disc space height is small, a slight change in height can result in large strains. The potential value of using strain to assess fusions can be most appreciated when rotation is less than two deg (Figure 5). Many studies have used two deg as a rotation threshold to classify a level as fused or not. (74–80) Figure 5 documents many levels where the rotation was < 2 deg yet the maximum strain was > 5%. If these were cases that were in fact not (yet) fused, then the reported fusion rates may have been less than those reported in some studies. Due to the lack of a validated gold standard, the true fusion status is not known.

**Figure 4:**
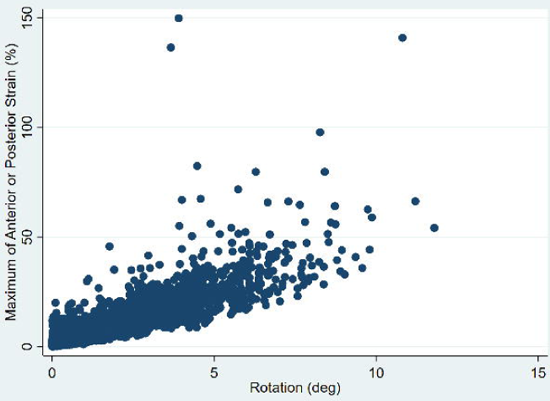
The maximum apparent strain at any point along the disc space was correlated with intervertebral rotation at levels treated using ACDF. Points are only plotted if there was at least 5 deg of rotation at one or more adjacent levels. 7624 data points are included in this graph.

**Figure 5:**
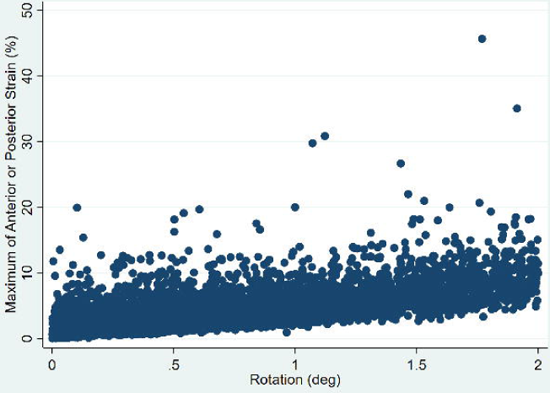
There are many post-ACDF levels where strain is > 2.5 % yet rotation is < 2 deg. These are from the same data as in Figure 4, but this figure only shows levels with rotation < 2 deg. There are 6139 data points plotted on this graph.

Further analysis of the pooled data revealed that with a rotation > 1 degree as the nonbridging criterion, a strain threshold of 4.8 was 88% sensitive and 85% specific to the rotation-based diagnosis. Strain- and rotation-based pseudoarthrosis diagnoses are generally consistent. Furthermore, a rotation of > 1 degree aligns with a > 5% strain in 86% of the measurements. Hence, discrepancies between the two assessment methods appear in approximately 14% of the patients. The implication is that in clinical practice, with 14 of every 100 diagnoses, only one of the approaches will correctly guide optimal clinical management. The challenge is to validate which is correct or how to combine them for a more accurate diagnosis.

In addition to the flexion-extension radiographs, a qualitative assessment of bridging bone was performed by a board-certified, fellowship-trained musculoskeletal radiologist at many of the time points in the pooled-data analysis. Some of the assessments were performed using CT, but most were performed from radiographs. In most of the studies, the radiologists had been trained to evaluate the flexion-extension radiographs. If rotation was above a certain level (typically three degrees), then the radiologist was trained to factor that into the assessment of bridging bone, with careful scrutiny of apparent bridging. Thus, bridging bone assessments are not always independent of intervertebral rotation. In no patients was a strain-based assessment used by the radiologist. Within that substantial limitation, a sensitivity/specificity analysis was completed to help understand the threshold level of strain (and rotation) that is most sensitive and specific to the radiologist assessment of no bridging. The optimal threshold of maximum strain (any point across the disc space) that optimizes sensitivity and specificity to the radiologist assessment of no bridging was 5.4%. At that threshold, the strain was 72% sensitive and 76% specific. Raising the threshold improved specificity (for no bridging) but worsened sensitivity. For example, at a threshold of 10%, the strain is 93% specific but only 50% sensitive due to a lack of bridging. At a threshold level of 3%, the strain is 86% sensitive but only 49% specific for a radiologist assessment of lack of bridging. The optimal threshold that can be used to predict meaningful clinical outcomes requires further research.

Notably, anterior- and posterior-most strains measured immediately adjacent to the superior and inferior levels initially increased and then gradually returned to near initial levels (Figure 6). These data are consistent with the hypothesis that strains at adjacent level discs initially increase due to the loss of motion at the fused level and with the hypothesis that the spine adapts to changes over time and that strains eventually return to more normal levels. These data suggest that strain measurements may help to diagnose changes in disc space properties other than for purposes of diagnosing pseudoarthrosis. Hypothetically, disc strain may be valuable for diagnosing the initial stages of disc degeneration and may therefore prove to be a companion diagnostic tool for biologic treatments for disc degeneration.

**Figure 6:**
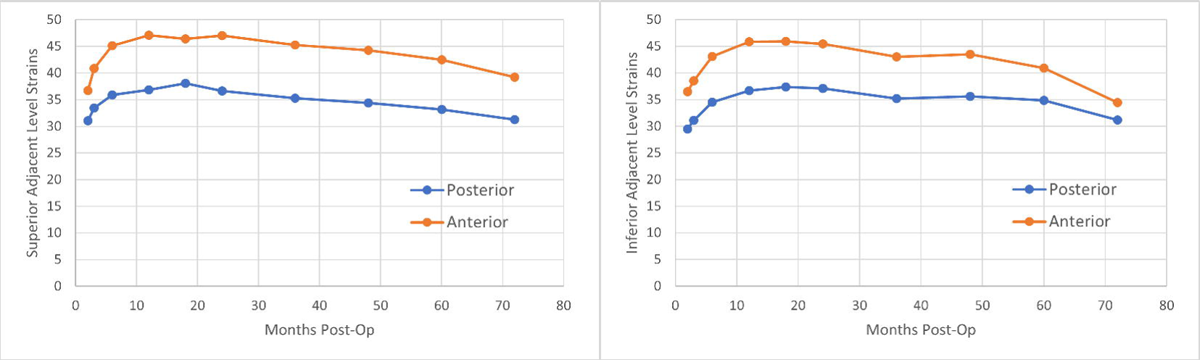
The change in average strain over time at the superior and inferior disc levels immediately adjacent to the spine fusion level(s).

## Discussion

Steps are proposed that may help progress toward a standardized diagnostic test for pseudoarthrosis. These criteria are based upon the following: the strain-based fusion assessment rationale; a proposed strain threshold based on both the failure strain of bone and expected measurement error; strains measured at unfused levels; the diagnostic performance of strain-based pseudoarthrosis diagnosis in a small study; and large sample size data on the reduction in strains that occur over time following ACDF surgery.

### Proposed steps toward standardizing the diagnosis of pseudoarthrosis

These proposed steps assume that intervertebral motion measurements are sufficiently reliable and assume that the goal of diagnosing pseudoarthrosis is defined by a lack of bridging bone:

- Quality control should be used to ensure that flexion-extension exams that stress the spine are sufficient to allow for reliable diagnosis of pseudoarthrosis.

- A flexion-extension protocol was used to encourage maximum flexion-extension effort while keeping the shoulders down to allow for analysis of lower cervical levels.
- If rotation is used, at least 5 deg of rotation is required at an adjacent, unfused level.
- Using a strain-based assessment requires at least 10% strain at an adjacent unfused level.
- With strain-based fusion assessment:

- A strain > 5% supported a diagnosis of a lack of bridging bone at a specific location. The 5% threshold is supported by laboratory data for the ultimate tensile strain of trabecular bone (81–83), in addition to tolerance for measurement error and by previously discussed retrospective data.
- Assess strain anteriorly and posteriorly to enable diagnosis of partial bridging that may progress across the disc space with time.
- With rotation-based fusion assessment:

- A rotation > 1 deg supports a general diagnosis of a lack of bridging bone(52), although there may be partial bridging at one aspect of the disc space.
- If there are significant differences in radiographic projection between flexion and extension, increase the threshold (e.g., rotation > 3 deg or strain > 8%), and accept that there will be diagnostic uncertainty.
- If available, image stabilization is used to support quantitative measurements, particularly when the rotation or strain are close to the proposed thresholds. Stabilization occurs when one vertebra is held in a constant position on the display as the flexion and extension views are alternately displayed. This approach facilitates visualization of relative motion between vertebrae, which can be subjectively interpreted. This approach may be particularly valuable for out-of-plane imaging.

Raising the thresholds (e.g., defining nonbridging as rotation > 2 deg or strain > 8%) will reduce the chances of incorrectly diagnosing genuinely fused cases as nonbridging. However, this approach will also increase the chances of mistakenly diagnosing cases that are not bridged as being fused. Youden’s index provides one option for determining a threshold that balances sensitivity and specificity.(84)

Both rotation- and strain-based diagnoses of pseudoarthrosis are easily calculated with automated vertebral landmarks. With high-quality imaging, both methods will likely agree when there is solid fusion or definitive intervertebral motion.

### Importance of an accurate diagnosis of pseudoarthrosis

There are several reasons why accurate diagnosis of pseudoarthrosis is important. First, a reliable diagnostic test would be useful for a substantial number of patients. More than 100K ACDF procedures are performed each year in the United States.(85) According to a large administrative database study, revision rates ranged from 9 to 11% in single and multilevel ACDF patients.(86) A 35% revision rate has been reported with multilevel ACDF.(87) Pseudoarthrosis can result in pain and poor outcomes in some patients.(88, 89) An accurate diagnosis of pseudoarthrosis is important in deciding whether to perform revision surgery.

Diagnosing a level that is actually “fused” as “not fused” can lead to unnecessary revision surgery and all associated costs: OR, anesthesia, hospital stay, postoperative therapy, possible need for postoperative orthosis or a bone growth stimulator, physical therapy, and postoperative surveillance imaging. The patient is also subjected to risks associated with further surgery, anesthesia, missed days of work, perioperative opioid pain management, and the stress of recovery. There is also a cost associated with preoperative work-up (such as preoperative CT scans, bone scans, dynamic radiographs, selective nerve or facet blocks, and laboratory testing). Furthermore, revising a fused segment that was presumed not to be fused does not adequately address the patient’s symptoms. When a level that is actually “not fused” is diagnosed as “fused,” the consequence can lead to costly nonoperative treatment modalities (such as pain management referrals, selective blocks, opioid pain management, and physical therapy), which do not address the actual underlying etiology of the patients’ symptoms. In some scenarios, this may lead patients to alternative palliative procedures, such as spinal cord stimulation for pain management, which is costly and leads to further surgical and implant-related morbidities. Adequate diagnosis of pseudoarthrosis is essential to avoid following either of these scenarios.

Accurate diagnosis of pseudoarthrosis at the appropriate clinical presentation is essential for treating patients with symptomatic pseudoarthrosis appropriately and in a timely manner to improve patient outcomes. Alternatively, in patients who have continued complaints following ACDF and have confirmed fusion, an accurate diagnosis of fusion allows clinicians to explore the alternative etiology of symptoms when further work is needed.

### Strain-versus rotation-based fusion assessment

Given that intervertebral rotation has long been a standard metric for assessing fusion status, it is valuable to explore whether a strain-based approach could enhance the accuracy of fusion assessment compared to rotation-based methods.

- Many different rotation thresholds have been used to classify a level as fused or not fused. (15, 55, 56) The optimal threshold can be determined only by experimental testing and requires a “gold-standard” method for determining true fusion status. This is difficult to accomplish with a large sample size. There is no biomechanically rational approach for predicting the optimal rotation threshold aside from the use of bone failure strain, and integrating endplate and disc height data is required to calculate the dependence of rotation on strain. This approach has no advantage over directly using a strain-based approach.
- Intervertebral rotation is a generic metric for the entire disc space and has no capacity to identify cases where, for example, there is anterior but not posterior bridging. Additional evidence is needed to determine whether location-specific bridging data are of clinical value. Hypothetically, this approach may help to justify a wait-and-see approach to symptomatic post-ACDF patients, where there is evidence that bridging may progress anteriorly to posteriorly.
- The stress and strain within a disc space vary based on the magnitude of rotation and the center of rotation (COR). For identical rotations, a COR near the disc’s center yields different stress and strain than when the COR is near the anterior side of the disc, as may occur with an anterior plate. While rotation-based evaluations do not account for the location of the COR, strain-based assessments do.

### The challenge of loading a spine sufficiently to apply strain-based fusion assessment

The strain-based fusion criterion could be most easily applied if a clinical test was developed that applied sufficient pure tension across the fusion surgery level such that the strain was much greater than 5% at every point across the disc space if the level was not fused. In that scenario, all points across the disc space could be reliably assessed for bridging. A lateral spine radiographic image in pure tension could be compared to an image with the spine in pure compression to calculate strain. However, a pure tension-compression protocol may not be clinically practical.

The images most commonly used to assess intervertebral motion (IVM) at the site of a spine fusion are radiographs of the patient in flexion and extension. As DeVine noted, “the data obtained are only as good as the amount of motion potentially generated by the method of flexion–extension imaging” (57). Several protocols for obtaining cervical spine flexion-extension radiographs have been described in at least some detail. (58–61) Patient instruction videos are available online for practical cervical spine flexion-extension protocols. (in English: https://youtu.be/AWooInuVo1Y; in Spanish: https://youtu.be/xYp1J7yvCgk) Additional validation of patient positioning protocols is needed.

Compressive loads can reduce the flexibility of the cervical spine.(58, 59) When upright, the combination of gravitational and muscular forces can stabilize the spine, potentially confounding the diagnosis of pseudoarthrosis. A patient positioning/loading scenario is necessary that allows for observable vertebral separation (well above 5% strain) if the level is not fused. Assessing whether the fusion surgery level is adequate for applying a strain-based (or rotation-based) fusion assessment is problematic. One solution is to calculate the strain across the disc space at adjacent non-fused levels. A strain that substantially surpasses the failure strain of bone across these disc spaces suggests that the adjacent level was adequately loaded, reinforcing confidence that the index level was also sufficiently loaded.

### Further research is essential

However, validating the clinical efficacy of diagnostic tests for pseudoarthrosis is challenging. Numerous studies have employed open surgical exploration as the gold standard against which alternative fusion assessment methods are compared, (23, 30, 31, 90, 91) even though there is no established validation of the accuracy and reproducibility of open surgical assessment. Other research has focused on the agreement between assessment methods and observers using specific techniques, (14) but these studies do not directly demonstrate the clinical value of a test.

An alternative approach could involve a randomized controlled trial that compares diagnostic tests, such as assessing using a CT exam versus strain-based fusion assessment from flexion–extension radiographs. The trial could include an outcome metric such as the NDI score at 6 months, one year, or two years following diagnosis. In settings where a CT exam is the current standard for diagnosis, an exploration could be conducted to determine whether strain-based assessment via flexion–extension radiographs offers sufficient diagnostic information, thereby avoiding the expenses and radiation exposure associated with a CT exam. Another option might involve evaluating the total cost of care (limited to cervical spine-related diagnosis and treatment codes) following the initial deployment of the diagnostic test, potentially using historical control data.

## Conclusion

The reliability of diagnosing pseudoarthrosis using traditional measures, including intervertebral rotation or interspinous process motion, along with subjective assessments of bone bridging, is notably low. Introducing a strain-based fusion assessment paradigm addresses some of these limitations and presents a potential pathway to achieving a reliable, standardized, and biomechanically explainable diagnosis of pseudoarthrosis, both in research studies and in clinical practice. Automated methods readily facilitate the acquisition of strain measurements.

To implement a strain-based (or rotation-based) fusion assessment, an imaging protocol must be established that can consistently stress the spine to the required extent. Clinical trials will be essential for evaluating the efficacy of a strain-based fusion assessment, discerning any potential advantages over alternative approaches, and validating clinical algorithms.

## Ethics considerations

This manuscript describes data obtained by retrospective automated analysis of previously collected spine radiographs. Pearl IRB (www.pearlirb.com) found the research to be Exempt according to 45 CFR 46.104(d)(4) Secondary Research Uses of Data or Specimens on 01/05/2024. IRB ID: 2024-0008

## Use of large language models

After the manuscript was drafted and reviewed by all the co-authors, ChatGPT (version 4) was used where a section of text was perceived to be deficient. The sections of text were input to ChatGPT with the request “Help make this text clearer and more concise.” When the ChatGPT response was perceived to improve without loss of accuracy or content, the ChatGPT response was integrated into the original text.

## Supplementary Material

Appendix 1: Review and critique of previous approaches for diagnosing pseudoarthrosis

Appendix 2: Disc strain error assessment using simulated radiographs

Brief videos summarizing some of the material in this paper are available at https://youtu.be/UJJTRKbDzI4, https://youtu.be/ptHdbRQm2Aw

## Data Availability

Data produced in the present study are available upon reasonable request to the authors

https://youtu.be/UJJTRKbDzI4

https://youtu.be/ptHdbRQm2Aw

