## Supplemental Appendix 1 for "Diagnosis of spine pseudoarthrosis based on the biomechanical properties of bone"

### Appendix 1: Review and critique of previous approaches to diagnosis of pseudoarthrosis

It is likely that radiographs or a CT exam will be reliable for fusion assessment when there is a definitive radiolucency at the fusion site, or if there is very definitive and solid bridging bone. However, the more common cases can be subtle and difficult to assess with suboptimal observer agreement. (1-4) The following pertains to the more subtle fusion assessments.

#### Issues with use of intervertebral rotation

Intervertebral rotation is a commonly used intervertebral motion criteria in spine fusion assessment. FDA guidelines suggest that with lumbar fusions, sagittal plane rotation over 5 deg indicates no fusion and under 5 deg indicates fusion (assuming that bridging bone is subjectively perceived bridging the disc space).(5) That high of a threshold may have been justified by un-assisted manual measurement error, but it is biomechanically nonsensical. Immediately after placing an interbody device and posterior screws and rods, the instrumentation alone will reduce rotation to under 5 deg. (6-8) If the disc space is packed with bone graft, it can be difficult to differentiate between bone graft and bridging bone until the graft is remodeled to oriented bone (9, 10). Thus, shortly after properly performed fusion surgery many levels would be classified as fused even though they are definitely not fused. An additional critical limitation of using intervertebral rotation to assess for motion at a fusion site is the dependence of rotation on the effort the patient exerts when asked to flex or extend. In large, multi-site clinical trials of patients that could be candidates for spine fusion surgery, preoperative rotation averages less than 5 degrees at the treatment level (11, 12), so based on the rotation component of the FDA recommended fusion criteria, many patients would be classified as fused before surgery based on intervertebral rotation using a > 5 deg threshold.

Fusion rates can be strongly influenced by the motion threshold selected(13). Bono et al and Gruskay et al review the various intervertebral rotation thresholds that have been used to classify a level as fused or not.(14) (15) Research has shown that residual motion can still occur in partially fused spinal levels. (14, 16-18) Finite element modeling revealed that intervertebral rotation can exceed 3 degrees with partial bridging bone formation.(16) For instance, if bridging bone forms only at the anterior part of the disc space, the posterior part may still open during flexion, indicated by widening of the spinous process. This can be detected by analyzing displacement at various points across the disc space; substantial displacement may occur posteriorly while no displacement is noted anteriorly at the location of bridging bone. Partial bridging is often considered a technical success, based on the expectation that stabilization will eventually lead to complete bridging. However, this assumption requires further research, particularly if the remaining intervertebral motion continues to cause symptoms despite the partial bridging.

#### Issues with use of spinous process widening

Spinous process widening (A.K.A. interspinous process motion) has been proposed by many investigators as a reliable method for cervical spine fusion assessment (see review by Oshina et al(19)). Note that if using spinous process widening to assess an ACDF, this is an indirect fusion assessment, since the surgery was not intended to fuse the spinous processes. There are three issues that compromise use of spinous process widening. The first is the challenge of reproducibly identifying the anatomic landmarks on the spinous processes that will be used to measure widening. There are no standards provided for this in routine clinical practice, spinous process can be highly variable between levels and between individuals, and with only the simple instruction, “place landmarks on the spinous processes and measure the change in spacing between flexion and extension”, it is unknown how reproducible the measurements would be among practicing clinicians. In an isolated and controlled research study, analysts can be carefully trained toward standardization. (20-23) That is much harder to accomplish across clinical medicine. Reliable spinous process widening assessment may be possible with machine learning, though that has yet to be developed. The second challenge, particularly with the cervical spine, is that the spinous process can be poorly visualized in the radiograph due to over-exposure or soft-tissues, or it may been removed or altered by laminectomy.(4) This can make it difficult or impossible to place landmarks on the spinous processes. The third challenge is from elastic deformation of the posterior elements.(24) Particularly with long slender spinous processes, due to the muscle forces required to position the neck in flexion or extension, the spinous processes can deform under load with respect to the vertebral body.(24) Anecdotally, this has been observed many times when viewing stabilized images from cervical flexion extension studies. Stabilization is the method where one selected vertebra is held in a constant position on a display as the flexion and extension images are alternately displayed.(25) This greatly facilitates interpretation of relative motion between vertebrae. Elastic deformation would most typically result in a successfully fused ACDF being classified as not fused due to measured interspinous process motion that is the result of elastic deformation of the posterior elements.

#### Translational motion

Some fusion criteria also use translational motion below a threshold to classify a level as fused (see review by Oshina et al(19)). No studies have provided data to allow an evidence-based understanding of how sagittal plane intervertebral translation could be useful independent of intervertebral rotation. Since most studies that included intervertebral motion as part of the fusion assessment criteria used flexion-extension radiographs to provoke intervertebral motion, intervertebral rotation would likely be the predominate motion, though that hypothesis has yet to be definitely tested.

#### Bridging bone assessment from CT

Definitive assessment of bridging bone from CT exams can be difficult due to the limited spatial resolution of clinical CT exams and the complex geometries of spine fusions.(26, 27) In research studies, high-resolution micro-CT exams can allow visualization of complex spine fusions(27-29), but this resolution is currently not available with clinical CT exams. Volume averaging and other technical limitations occur with CT exams, and that prevents the spatial resolution required to definitively visualize geometrically irregular spicules of bridging bone or thin non-mineralized layers that can occur in cases of pseudoarthrosis (26, 30-32). Trabeculae are typically under 0.2 mm thick (33). The thinnest CT slices available in clinical practice are typically > 0.625 mm, and even the in-plane resolution is too low to discern individual trabeculae. Each picture element (pixel) in each axial slice of a CT exam (and orthogonal slices reconstructed from the axial slices) average together all tissue within the thickness of the slice and area of the pixel. If the tissue at any specific location in the axial slice is predominately bone, it can be perceived as part of a bone bridge, even if there is actually a thin cleft through the fusion site that can be seen with histology or micro-CT(30, 34, 35). In addition, artifacts around metal hardware can further confound assessment of bridging bone(10, 36, 37). Early in the post-surgery period, bone graft must be remodeled into bridging bone, and it is difficult to discern the difference between tightly packed but not fused bone graft material, versus remodeled and bridged bone. Finally immature woven bone that has formed a bridge across the disc space can provide substantial stiffness to the treated level despite not appearing radiographically as a solid fusion.(38)
