## Supplemental Appendix 2 for "Diagnosis of spine pseudoarthrosis based on the biomechanical properties of bone"

### Appendix 2: Disc strain error assessment using simulated radiographs

#### Methods

To assess errors in disc strain measurements from point-source radiographs, simulated Digitally Reconstructed Radiographs (DRR) were used. This was done to eliminate errors due to the challenges of placing landmarks on two dimensional projections of the spine. Nine fully anonymized thin slice CT exams of the cervical spine were used to create the DRRs. Landmarks were placed in 3-D on the anterior and posterior most corners of the superior and inferior and plates in the mid-sagittal plane using the 3-D Slicer image computing platform(slicer.org, last accessed Nov 2023).[1] To enable quantification of the amount of out-of-plane error in the radiographs, landmarks were also placed in the mid-coronal plane of each vertebral body on the left and right most aspects of the inferior and superior table endplates. Mid-coronal plane landmark placement was standardized to the extent possible, though this is difficult due to the variability of uncovertebral joints and the complex geometry of cervical endplates. Metal balls were virtually added to the CT exams at each landmark location prior to generating the DRRs. These landmarks, visible as points along the endplate anatomy in the simulated radiographs, helped measure the extent of X-ray beam misalignment with the endplates and the posterior wall of the vertebra. (Figure 1) The extent of misalignment, or "out-of-plane" error was measured in both the cranial-caudal and anterior-posterior directions.

Custom code was used to create digitally reconstructed radiographs of each spine. This code made calls to Plastimatch DRR (http://plastimatch.org) to generate the simulated radiographs from the 3-D CT exams. The projection matrices specified by Plastimatch DRR were reproduced in the code to precisely calculate the two-dimensional coordinates of every landmark as projected onto each 2-D radiograph. The source-to-image distance was set at 70 inches and the distance between image and mid-sagittal plane of the spine was set to ½ of a typical shoulder width. The first simulated radiograph was centered on the C3-C4 disc space. An additional 40 radiographs of each spine were created where the X-ray beam was randomly tilted between ±10 deg in the coronal and axial planes, where the beam center was randomly moved ±2 endplate widths from the initial beam center, and where the X-ray was rotated ±25° about the image center. These were intended to represent typical clinical variability in obtaining lateral spine flexion-extension radiographs.

Two-hundred randomly selected pairs of simulated radiographs from each spine were then used to represent flexion and extension radiographs. Intervertebral rotation, translation, and disc strains were calculated using SpineCAMP™ (Medical Metrics, Inc, Houston, TX). Disc strains were calculated at 11 equally spaced points across the disc space from anterior to posterior (see Figure 1 of the paper referencing this appendix), and the maximum disc strain was used for the analysis of errors. In this experiment, there should be no intervertebral motion or disc strain measured from the paired radiographs. Any IVM or strain that is measured from the projected landmarks is due to error that occurs when these measurements are made from radiographs. The amount of cranial-caudal and anterior-posterior out-of-plane was measured (Figure 1) for each disc space as the average of the four out-of-plane measurements at the inferior and superior endplate of the inferior and inferior vertebrae. 9000 levels were analyzed (9 spines x 200 pairs x 5 disc levels per spine). The mismatch in the out-of-plane measurements between the paired images was calculated for each level. An example of high cranial-caudal mismatch is provided in Figure 2.

### Results

Recalling that any measured intervertebral motion or disc strain is due to the error that can occur when making these measurements from radiographs, measured disc strains averaged 2.7 ±3.4. Based on multivariate analysis of variance, the variables that primarily influence the measured disc strains were the disc height and the amount of cranial-caudal out-of-plane mismatch. The error in disc strain increases with narrow disc spaces and with higher mismatch between the cranial-caudal out-of-plane in the two images. The mismatch between images in radiographic projection did not only effect the disc strains; intervertebral rotations measured as high as 1.9 deg and translations measured as high as 4.2 % endplate width.

### Discussion

The potential error in measuring disc strains from radiographs can be conceptualized using a rectangle, which simplistically represents the disc space as viewed in the mid-sagittal plane. Consider a rectangle hanging a distance from a wall and illuminated by a movable light source. As the light source shifts vertically or horizontally, the rectangle's shadow distorts, altering its perceived height and width. If only the shadow of the rectangle is available for analysis and the light source's position is unknown, changes in the rectangle's dimensions between two images might be incorrectly attributed to changes in the rectangle's shape, rather than light positioning. Similarly, without precise knowledge of the X-ray source's position relative to the spine, apparent changes in disc space height due to varied radiographic projection may be misinterpreted as strain, even when no actual strain exists.

The experiment with simulated radiographs helps to quantify the errors that can be expected in disc strain measurements made from radiographs with variable radiographic projection. This experiment is most relevant to the challenge of determining if a level is solidly fused such that there is no actual measurable intervertebral motion. The experiment supports that if the radiographic projection of the disc space is similar between flexion and extension, then measured strain due to no more than moderate differences in radiographic projection would average < 3% and that would be consistent with solid fusion. However, if the radiographic projections differ substantially, measured strains could be as high as 10% even though the level is solidly fused.

It is not just strains that are affected by variable radiographic projection. Intervertebral rotations as high as 1.9 deg and translations as high as 4.2 % endplate width were measured due only to differences in radiographic projection between images. If the radiographic projections in the two images are very similar, intervertebral rotation and translation errors can be expected to be very small, since for the 9000 levels analyzed, rotations averaged 0±0.3 deg, and translations averaged 0±0.8 % endplate width. However, as the mismatch in the amount of out-of-plane between images becomes large, rotation and translation must be interpreted with higher error limits.

Solutions to the problem of variable radiographic projection can include: quality controlled flexion-extension radiographs using a validated protocol (e.g., https://youtu.be/AWooInuVo1Y) to minimize out-of-plane errors; a neural network trained to quantify the amount of out-of-plane error so that the measurement confidence intervals can be provided; neural networks trained to create a perfectly aligned radiograph from a mis-aligned radiograph. In addition to accounting for variable radiographic projection when interpreting disc strain measurements, disc height also must be considered, since the errors due to radiographic projection variability are magnified when the disc space is very narrow. That is a consequence of apparent disc tensile strain being calculated as the change in disc height divided by the smallest disc height, so when the disc height is very small, the ratio becomes unstable.

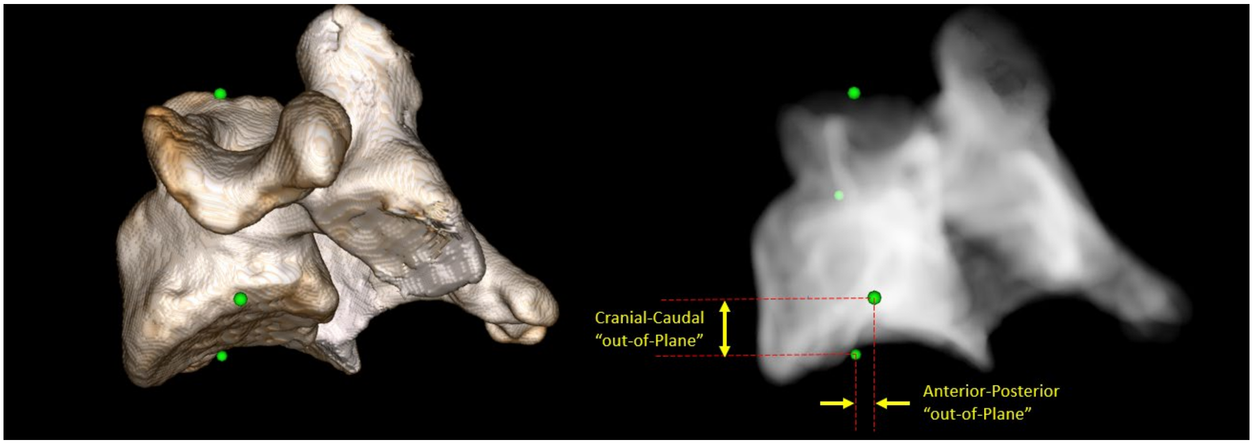


Figure 1: The figure displays a 3D representation of a C4 vertebra (right) with markers on the left and right sides of the superior and inferior endplates, that were used to quantify the "out-of-plane" error or the degree of vertebral misalignment with the X-ray beam. On the right, is the vertebra as it appears on a simulated radiograph, highlighting the measurement protocol for cranial-caudal and anterior-posterior misalignment. These measurements, made on both the superior and inferior endplates of the vertebrae above and below the disc space, are averaged to obtain a single cranial-caudal and a single anterior-posterior out-of-plane error for each disc level. Measurements are in units of %AP endplate width.

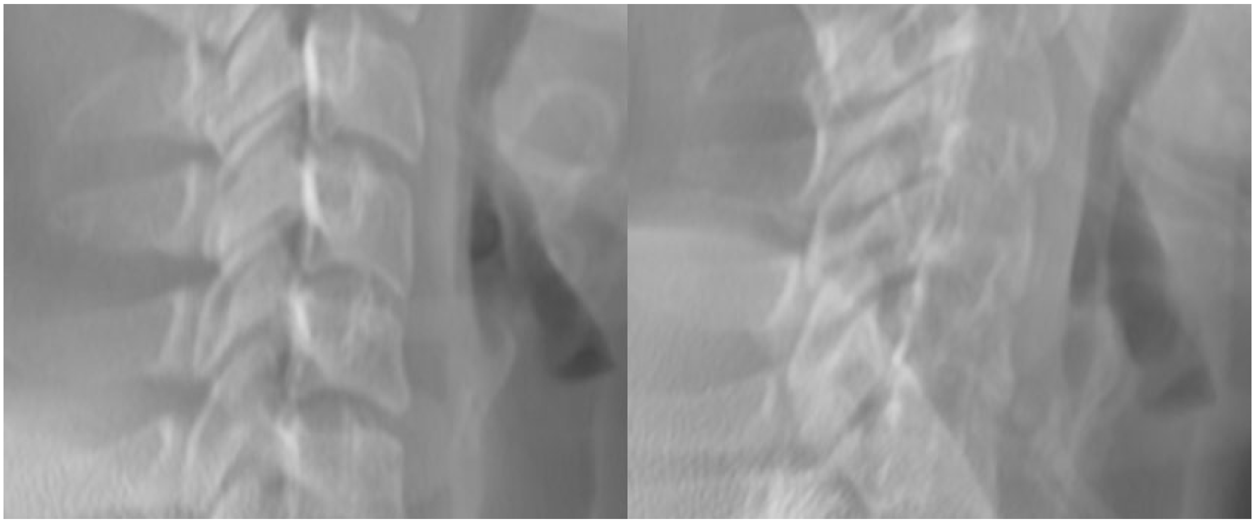

Figure 2: An example of paired simulated radiographs with a high (25% endplate width) mismatch in the amount of cranial-caudal out-of-plane at the C4C5 disc space (centered in the images).
